# Incidence of Severe Acute Respiratory Syndrome Coronavirus-2 infection among previously infected or vaccinated employees

**DOI:** 10.1101/2021.07.03.21259976

**Authors:** N Kojima, A Roshani, M Brobeck, A Baca, JD Klausner

**Affiliations:** Department of Medicine, University of California Los Angeles, Los Angeles, 90095; Curative Inc., San Dimas, CA; Department of Population and Public Health Sciences, University of Southern California, Keck School of Medicine, Los Angeles, 90033

**Keywords:** COVID-19, SARS-CoV-2, Incidence, Reinfection, Prior Infection, Vaccination

## Abstract

**Introduction:** The protective effect of previous infection versus vaccination is poorly studied. Among a clinical laboratory that has been conducting routine workforce screening since the beginning of the pandemic, we aimed to assess the relative risk of Severe Acute Respiratory Syndrome Coronavirus-2 (SARS-CoV-2) infection among individuals who were SARS-CoV-2 naïve, previously infected, or vaccinated.

**Methods:** Using an electronic laboratory information system, employees were divided into three groups: (1) SARS-CoV-2 naïve and unvaccinated, (2) previous SARS-CoV-2 infection, and (3) vaccinated. Person-days were measured from the date of the employee first test and truncated at the end of the observation period. SARS-CoV-2 infection was defined as two positive SARS-CoV-2 PCR tests in a 30-day period. Individuals with fewer than 14 days of follow up were excluded. Incidence estimates and the 95% confidence intervals were calculated using the Poisson Exact equation. The incidence rate ratio (IRR) was used as a measure of association between groups. Analyses were performed on StataSE (StataCorp, College Station, TX).

**Results:** We identified 4313, 254 and 739 employee records for groups 1, 2, and 3, respectively. The median age of employees was 29.0 years (interquartile range: 23.6, 39.9). During the observation period, 254, 0, and 4 infections were identified among groups 1, 2, and 3, respectively. Group 1 had an incidence of 25.9 per 100 person-years (95% CI: 22.8-29.3). Group 2 had an incidence of 0 per 100 person-years (95% CI: 0-5.0). Group 3 had an incidence of 1.6 per 100 person-years (95% CI: 0.04-4.2). The IRR of reinfection among those with previous infection compared to SARS-CoV-2 naïve was 0 (95% CI: 0-0.19). The IRR of those vaccinated compared to SARS-CoV-2 naïve was 0.06 (95% CI: 0.02-0.16). The IRR of those vaccinated compared to prior SARS-CoV-2 was 0 (95% CI: 0-4.98).

**Conclusion:** Previous SARS-CoV-2 infection and vaccination for SARS-CoV-2 were associated with decreased risk for infection or re-infection with SARS-CoV-2 in a routinely screened workforce. The was no difference in the infection incidence between vaccinated individuals and individuals with previous infection. Further research is needed to determine whether our results are consistent with the emergence of new SARS-CoV-2 variants.

## Introduction

Prior reports have found lower rates of Severe Acute Respiratory Syndrome Coronavirus-2 (SARS-CoV-2) infections among those with prior infection or vaccination.^1, 2^ Although an association between vaccination and reduction of SARS-CoV-2 incidence has been well described, how the incidence among individuals with previous infection compares to vaccinated individuals remains unclear.

## Methods

In March 2020, Curative, a SARS-CoV-2 testing company, began routinely screening its workforce with an Food and Drug Administration-authorized SARS-CoV-2 polymerase chain reaction (PCR)-based test.^3^ The workforce was screened daily. A standardized employee testing database was implemented on 8 May 2020. On December 15, 2020, vaccination with either the BNT162b2 or mRNA-1273 vaccines became available. Routine screening has continued through July 2021.

The SARS-CoV-2 naïve, unvaccinated group was defined as any employee without previous infection that tested from 8 May up to 15 December 2020 (when vaccination became available). The previously infected, unvaccinated group was defined as any employee with documented previous SARS-CoV-2 infection (at least 2 positive PCR tests) between 8 May to 15 December 2020. The vaccinated group was defined as any employee with documented completion of vaccination through 1 July 2021.

Person-days were measured from the first test date to last test date up to December 15^th^, 2020 for groups 1 and 2 and up to July 1, 2021 for group 3. We defined SARS-CoV-2 infection as two positive PCR tests in a 30-day period. Individuals with fewer than 14 days of follow up were excluded. Incidence in 100 person-years with 95% confidence intervals (95% CIs) was calculated with the Poisson Exact equation. The incidence rate ratio (IRR), the ratio of confirmed COVID-19 cases per 100 person-years of follow up with 95% CIs, was used as a measure of association between groups. Analyses were performed on StataSE (StataCorp, College Station, TX). The study of de-identified electronic medical record data was determined by the Advarra institutional review board (Pro00054560) to be exempt from review.

## Results

We identified 4313, 254 and 739 employee records for the naïve and unvaccinated group (Group 1), the previously infected and unvaccinated group (Group 2) and the vaccinated without previous infection group (Group 3), respectively. The median age of employees was 29.0 years (interquartile range: 23.6, 39.9). During the observation period, 254, 0, and 4 SARS-CoV-2 incident infections were identified among Groups 1, 2, and 3, respectively. (Table).

**Table.**
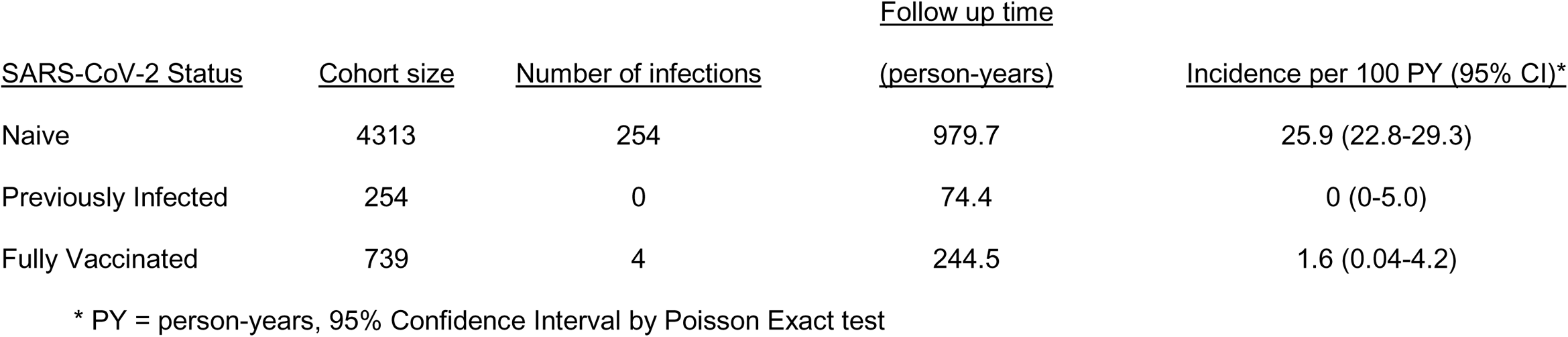
Incidence of SARS-CoV-2 infection among employees SARS-CoV-2 naïve, previously infected or vaccinated in a clinical laboratory workforce, 2020-2021

| <u>SARS-CoV-2 Status</u> | <u>Cohort size</u> | <u>Number of infections</u> | <u>Follow up time</u><br><u>(person-years)</u> | <u>Incidence per 100 PY (95% CI)*</u> |
| --- | --- | --- | --- | --- |
| Naive | 4313 | 254 | 979.7 | 25.9 (22.8-29.3) |
| Previously Infected | 254 | 0 | 74.4 | 0 (0-5.0) |
| Fully Vaccinated | 739 | 4 | 244.5 | 1.6 (0.04-4.2) |

\* PY = person-years, 95% Confidence Interval by Poisson Exact test

The naïve, unvaccinated group had a SARS-CoV-2 incidence of 25.9 per 100 person-years (95% CI: 22.8-29.3). The previously infected, unvaccinated group had an incidence of 0 per 100 person-years (95% CI: 0-5.0). The vaccinated group had an incidence of 1.6 per 100 person-years (95% CI: 0.04-4.2). The IRR of reinfection among those with previous infection compared to those SARS-CoV-2 naïve was 0 (95% CI: 0-0.19). The IRR of those vaccinated compared to those SARS-CoV-2 naïve was 0.06 (95% CI: 0.02-0.16). The IRR of those vaccinated compared to those previously SARS-CoV-2 infected was 0 (95% CI: 0-4.98).

## Discussion

In the workplace setting, we observed a lower incidence of SARS-CoV-2 infection among those with previous SARS-CoV-2 infection or SARS-CoV-2 vaccination with either the BNT162b2 or mRNA-1273 vaccines. Either prior infection or vaccination was associated with a dramatic decreased risk for infection or re-infection with SARS-CoV-2. The was no difference in the incidence of SARS-CoV-2 infection or re-infection between individuals who were vaccinated and individuals with prior SARS-CoV-2 infection, respectively.

Our findings are similar to other studies that compared the incidence of SARS-CoV-2 infection among those with prior SARS-CoV-2 infection and vaccination to unvaccinated antibody seronegative individuals. In a study conducted in Oxfordshire, UK, researchers reported that they found no differences in immunity induced by natural infection and vaccination with the BNT162b2 or ChAdOx1 nCOV-19 vaccines among a cohort of 13,109 healthcare workers.^4^ Another group of researchers studying a group of 52,238 employees of the Cleveland Clinic Health System found that those with previous SARS-CoV-2 infection and those who were vaccinated had lower rates of SARS-CoV-2 infection compared to those who were SARS-CoV-2 naïve and unvaccinated.^5^

After vaccination or natural infection, many mechanisms of immunity exist including humoral and cellular immunity.^6-8^ It is known that SARS-CoV-2 infection induces specific and durable T cell immunity against multiple SARS-CoV-2 spike (S) protein targets (or epitopes) as well recognition of other SARS-CoV-2 proteins. The broad diversity of T-cell viral recognition serves to enhance protection to SARS-CoV-2 variants,^7^ with recognition of at least three SARS-CoV-2 variants (B.1.1.7 [U.K.], B.1.351 [South Africa], and B.1.1.248 [Brazil]).^9^ Additionally, a memory B cell response to SARS-CoV-2 evolves between 1.3 and 6.2 months after infection that is consistent with immune persistence.^10^

Our findings were limited by the observational nature of the study. It is possible, but unlikely, that employees could have tested positive outside of the employee testing program. In addition, because allocation to each exposure group was not random, there might be differences between groups in the risk of repeat exposure over time. The study was strengthened by the high incidence among those naïve and unvaccinated, the large sample size and large number of person-years of follow up in each group.

## Conclusion

We found a strong association between prior SARS-CoV-2 infection and vaccination for SARS-CoV-2 with either BNT162b2 or mRNA-1273 and the reduced incidence of SARS-CoV-2 when compared to those naïve and unvaccinated to SARS-CoV-2. The was no difference in the incidence of SARS-CoV-2 between individuals who were vaccinated and individuals with prior SARS-CoV-2 infection. Combined with prior studies, our findings should provide increased confidence that those previously infected are at very low risk for repeat infection. Further research is needed to determine whether our results are consistent with the emergence of new SARS-CoV-2 variants.

## Data Availability

Data is available to requesting parties.

## Acknowledgements

The staff at Curative Inc.

